# Development and Evaluation of MADDIE: Method to Acquire Delivery Date Information from Electronic Health Records

**DOI:** 10.1101/2020.07.30.20165381

**Authors:** Silvia P. Canelón, Heather H. Burris, Lisa D. Levine, Mary Regina Boland

## Abstract

**Objective:** To develop an algorithm that infers patient delivery dates (PDDs) and delivery-specific details from Electronic Health Records (EHRs) with high accuracy.

**Materials and Methods:** We obtained EHR data from 1,060,100 female patients treated at Penn Medicine hospitals or outpatient clinics between 2010-2017. We developed an algorithm called MADDIE: Method to Acquire Delivery Date Information from Electronic Health Records that infers a PDD for distinct deliveries based on EHR encounter dates assigned a delivery code, the frequency of code usage, and the time differential between code assignments. We validated MADDIE’s PDDs against a birth log independently maintained by the Department of Obstetrics and Gynecology.

**Results:** MADDIE identified 50,560 patients having 63,334 distinct deliveries. MADDIE was 98.6% accurate (F_1_-score 92.1%) when compared to the birth log. The PDD was on average 0.68 days earlier than the true delivery date for patients with only one delivery (± 1.43 days) and 0.52 days earlier for patients with more than one delivery episode (± 1.11 days).

**Discussion:** MADDIE is the first algorithm to successfully infer PDD information using only structured delivery codes and identify multiple deliveries per patient. MADDIE is also the first to validate the accuracy of the PDD using an external gold standard of known delivery dates as opposed to manual chart review of a sample.

**Conclusion:** MADDIE infers delivery dates and delivery-specific details from the EHR with high accuracy and relies only on structured EHR elements while harnessing temporal information and the frequency of code usage to identify accurate PDDs.

## 1. BACKGROUND AND SIGNIFICANCE

Traditional research studies often suffer from a lack of racial and ethnicity diversity,[1] in part because of the complexity of recruiting diverse populations.[2,3] An approach that can enable research in diverse populations involves the use of existing clinical records repurposed for research. Since the passage of the Affordable Care Act, many hospitals, academic medical centers and outpatient clinics have transformed their previously paper-based clinical records to electronic records.[4] These Electronic Health Records (EHRs) can be repurposed for research if appropriate biomedical informatics methods and algorithms are developed and applied to ensure the data are of sufficient quality.[5]

EHR data is increasingly being used for population-based studies for health status surveillance,[6] identification of patterns in patient outcomes,[7,8] quality and operations improvement, [9,10] risk assessment and prediction, [11,12] among other translational and clinical research objectives.[13] Secondary use of EHRs can be particularly helpful when considering patient populations that are typically underrepresented in clinical trials designed to explore strategies, treatments, or devices that could be effective in preventing or treating disease. Under-represented populations in research include pregnant individuals and elderly patients,[14] as well as ethnic, racial and other minorities. [15]

One of the challenges of using EHR data in research is that it contains a combination of inpatient and outpatient records with diagnosis and procedure codes, and encounters, which chronicle a patient’s medical history without necessarily containing detailed information about a particular pregnancy. Researchers interested in studying pregnancy and delivery-related outcomes, have developed various methods to extract structured information on pregnancies from the EHR. Methods that have been developed to infer delivery episodes and delivery dates from the HER [16–22] and/or claims[18,19,21,23] databases have used a variety of classification code systems. The *International Classification of Diseases* (ICD) coding system is the most widely used and exists as two modified versions in the United States: the *9*^*th*^*Revision* (ICD-9 code set used before October 2015, and the *10*^*th*^ *Revision* (ICD-10) used afterwards.[24] Both the clinical modification (CM) and procedural coding systems (PCS) of these coding systems are used primarily for billing and insurance reimbursement purposes but can also serve as a proxy for a patient’s pregnancy state when linked to delivery outcomes or other pregnancy-related data. Researchers have also used EHR data to evaluate post-market approval assessment of medication safety in pregnancy [21,22,25] and EHR data has the potential to facilitate investigation of other prenatal exposures on pregnancy-related outcomes.[26] The former have mainly utilized algorithms to estimate delivery outcomes rather than the date of delivery,[21,22,25] and the latter to estimate fetal loss utilizing a database with built-in mother-infant linkages.[26]

Accuracy in inferring delivery episodes and delivery dates is critical in assessing epidemiological outcomes and can be optimized using an external form of validation. Some studies estimating gestational age have used the date of the last menstrual period, ultrasound information, and/or clinical estimates of the gestational age as validation [16,17] and others have relied on manual chart review.[20] In addition, research studies have extracted delivery outcomes from medical record databases and validated the findings using independent records, [22] laboratory tests, [27] manual review,[18–20] or by comparing with national estimates of outcome rates.[21,23] Previously published delivery date estimation methods have relied on manual review by trained abstractors of a random sample of the data due to the labor-intensive requirements of the task,[19,20] highlighting a major limitation and opportunity for improvement (**Table 1**). In general, these prior manual review approaches focus on clinician evaluation of timelines of structured pregnancy-related codes to assess whether or not, and when, a pregnancy has occurred A subset of prior studies that use large clinical records systems to investigate perinatal health outcomes relied upon linking maternal and child records, either through insurance information [28] or EHR-maintained mother-infant links.[29] Not all institutions, Penn Medicine included, have the ability to link historical maternal and child records highlighting another need for a robust and validated delivery identification algorithm that captures a patient’s date of delivery.

**Table 1.**
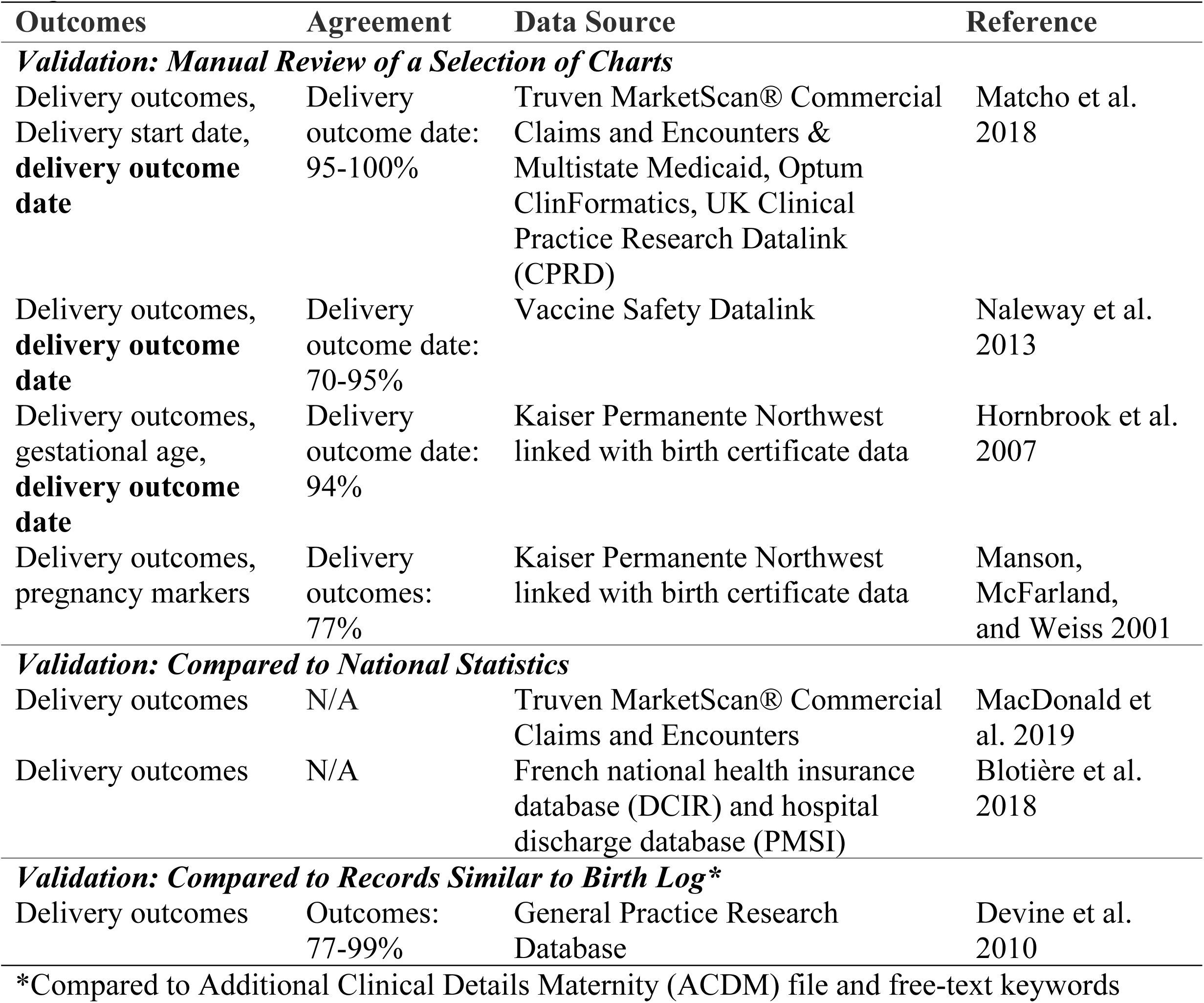
Details of Validation Methods Used by Previously Published Delivery-Detection Algorithms.

| Outcomes | Agreement | Data Source | Reference |
| --- | --- | --- | --- |
| ***Validation: Manual Review of a Selection of Charts*** |  |  |  |
| Delivery outcomes,<br>Delivery start date,<br>**delivery outcome date** | Delivery<br>outcome date:<br>95-100% | Truven MarketScan® Commercial Claims and Encounters & Multistate Medicaid, Optum ClinFormatics, UK Clinical Practice Research Datalink (CPRD) | Matcho et al. 2018 |
| Delivery outcomes,<br>**delivery outcome date** | Delivery<br>outcome date:<br>70-95% | Vaccine Safety Datalink | Naleway et al. 2013 |
| Delivery outcomes,<br>gestational age,<br>**delivery outcome date** | Delivery<br>outcome date:<br>94% | Kaiser Permanente Northwest linked with birth certificate data | Hornbrook et al. 2007 |
| Delivery outcomes,<br>pregnancy markers | Delivery<br>outcomes:<br>77% | Kaiser Permanente Northwest linked with birth certificate data | Manson, McFarland, and Weiss 2001 |
| ***Validation: Compared to National Statistics*** |  |  |  |
| Delivery outcomes | N/A | Truven MarketScan® Commercial Claims and Encounters | MacDonald et al. 2019 |
| Delivery outcomes | N/A | French national health insurance database (DCIR) and hospital discharge database (PMSI) | Blotière et al. 2018 |
| ***Validation: Compared to Records Similar to Birth Log\**** |  |  |  |
| Delivery outcomes | Outcomes:<br>77-99% | General Practice Research Database | Devine et al. 2010 |

\*Compared to Additional Clinical Details Maternity (ACDM) file and free-text keywords

The objective of our study is to develop a delivery date algorithm designed to 1) accurately identify women who delivered and 2) determine their date of delivery with an algorithm using structured EHR data alone. We also validate our algorithm against an independently maintained departmental log of birth records used to provide vital statistics to the state, available for one of our six hospitals within the Penn Medicine health system.

## 2. MATERIALS AND METHODS

We developed an algorithm called **MADDIE**: **M**ethod to **A**cquire **D**elivery **D**ate **I**nformation from **E**lectronic Health Records that infers each Patient Delivery Date (PDD) from a patient’s entire timeline of structured EHR data that includes a combination of inpatient and outpatient data from at least 1,048 clinics.[30] We developed MADDIE and validated against an independently maintained birth log containing detailed information regarding the deliveries from one of the Penn Medicine hospitals (**Figure 1**).

**Figure 1.**
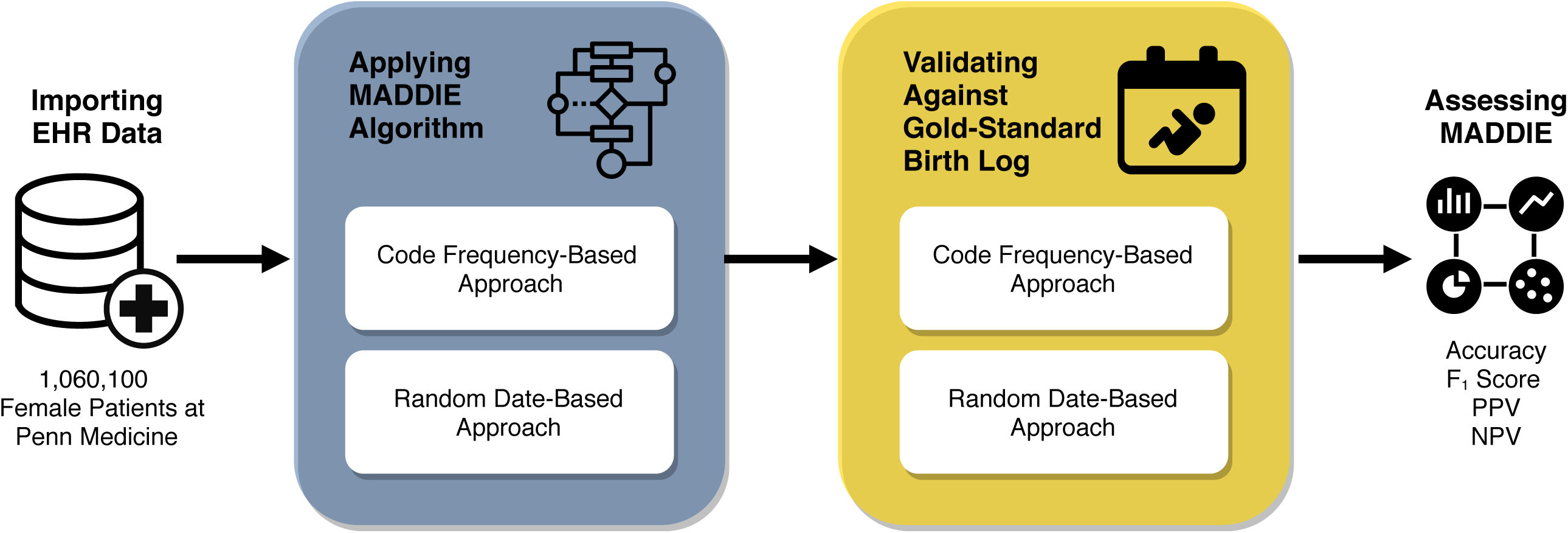
Process pipeline for the approach taken in this study.

Overall, MADDIE uses codes and their frequency of usage coupled with temporal information regarding the timing of code assignments to infer each PDD. MADDIE uses EHR encounter dates with an assigned delivery code along with their temporal sequence to identify a PDD.

### 2.1 Delivery Episode Algorithm

#### 2.1.1 Our Cohort: All Female Patients Treated at Penn Medicine

We obtained EHR data for 1,060,100 female patients with visits to inpatient or outpatient clinics within the Penn Medicine health system between 2010 and 2017. No restrictions were made to the patient population (i.e. reproductive age) except that the patients were 18 years old at time of data extraction in 2017-2018. The purpose of developing the MADDIE algorithm was to uncover each patient’s deliveries (i.e., instances where they gave birth) with the understanding that these should occur during the typical age of reproductive potential in females (<60 years of age). However, we did not exclude older patients, but applied our algorithm on the entire cohort. We used U.S.-modified ICD version 9 (ICD-9) and version 10 (ICD-10) codes (**Supplement File 1**) to identify 50,560 patients with delivery diagnoses or delivery procedures during any inpatient or outpatient clinic visit (**Figure 2**). While most deliveries are recorded during inpatient clinic visits, data from outpatient clinic visits was included in order to capture deliveries that start in outpatient clinics and transition to inpatient care as the delivery progresses. We anticipated that this would capture the entire delivery episode more completely than if outpatient encounters were excluded.

**Figure 2.**
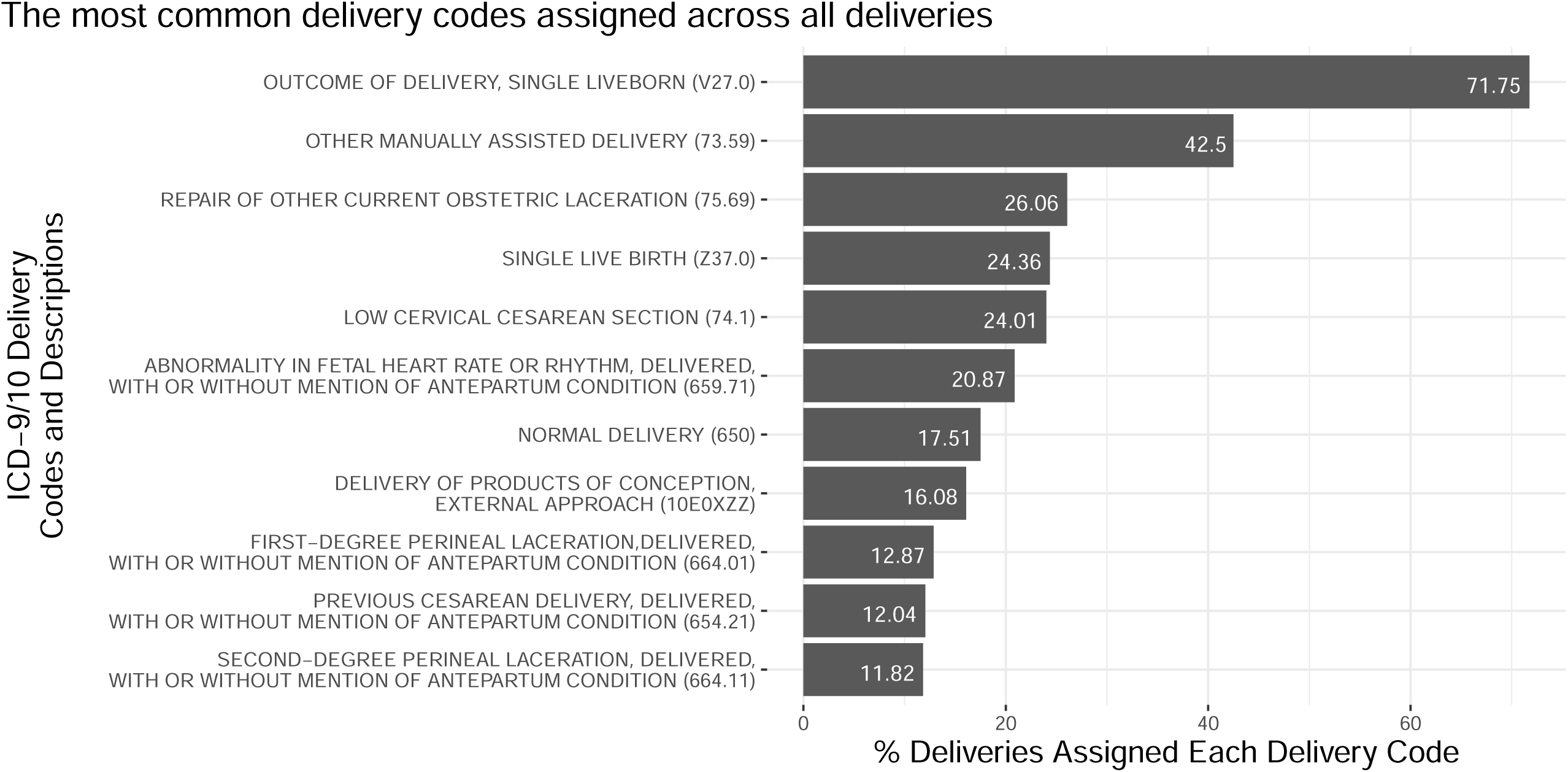
The distribution of delivery-related codes assigned to patient encounters and used to identify distinct delivery episodes for each patient.

Identifying patients with delivery diagnoses was only the first step. We next needed to separate out each delivery within each patient’s record. This is especially important as many of our patients have more than one delivery within the Penn Medicine health system. To identify the PDD for patients with only one encounter date having one assigned delivery code was fairly straightforward. We designed our algorithm, MADDIE to handle more complex cases involving more than one delivery code assigned over a series of encounter dates by grouping those codes into a “delivery episode”. MADDIE requires as input only the unique patient identifier or Medical Record Number (MRN), ICD codes, and encounter dates corresponding to relevant codes. MADDIE then separates the data into delivery episodes when appropriate and computes the frequency of code usage to accurately infer the PDD. More details are contained in the sections below.

#### 2.1.2 Distinguishing Separate Deliveries and Defining the ‘Delivery Episode’

For patients who had only one encounter date where delivery codes were assigned, then MADDIE assigned that date as the PDD. If a patient had multiple encounter dates with delivery codes, the dates were sorted in chronological order and the difference in days between each consecutive encounter date was calculated. We assumed that a gap of 180 days between delivery-related and coded visits could be considered as part of two separate deliveries. If a patient became pregnant soon after their first delivery in our records, we are ensuring that their second delivery occurs at least 24 weeks later, which should hold true for both term and most preterm births [31,32] (**Figure 3**). We did not consider fetal loss or miscarriages in this algorithm, but instead focused only on completed pregnancies with delivery. This includes stillbirth (as a ‘delivery’ took place), but not fetal loss.

**Figure 3.**
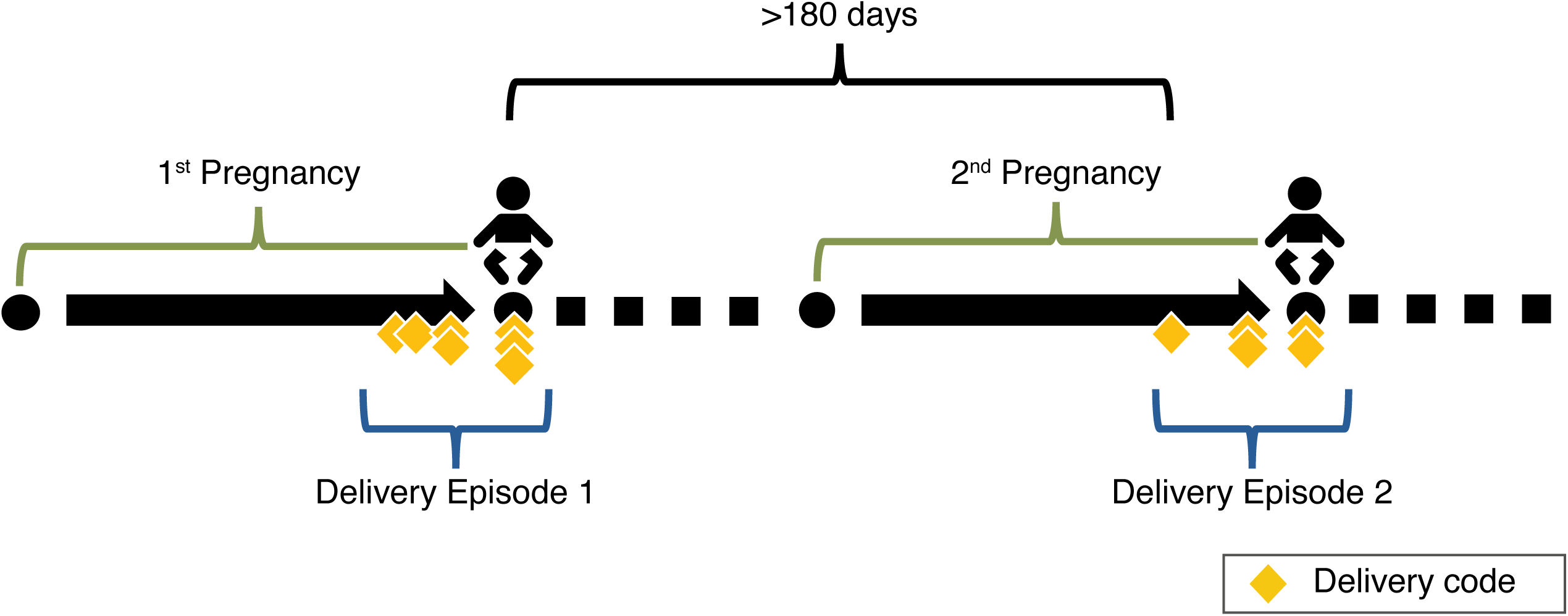
Schematic depicting a patient’s pregnancy timeline within the EHR, including billing code instances for delivery.

#### 2.1.3 Identifying Delivery Episodes

If the difference in days between the ordered delivery encounters was less than or equal to 180 days, both dates were considered part of the same delivery episode. Once the difference between encounter dates was greater than or equal to 180 days, the delivery episode counter increased to mark the beginning of a new delivery episode. This could occur multiple times for a single patient and thereby allow them to have multiple different deliveries and delivery episodes, provided those records were available within our dataset.

#### 2.1.4 Selecting the Delivery Date

In the event that a delivery episode included more than one delivery encounter date, MADDIE used a code frequency-based approach to select the delivery date. The code frequency-based approach assumed that the encounter date assigned the most delivery codes was the most likely to be the delivery date, and MADDIE selected this date as the PDD. We ignored the timestamp (e.g., 8:33 a.m.) but focused only at the day level (e.g., 04-01-2020). Therefore, the date with the greatest number of delivery codes assigned was considered the PDD. If two or more encounter dates had the same frequency of delivery codes, the most recent (i.e., the latest) encounter date was selected as the PDD. This decision was made assuming the delivery encounters were part of a sequence of delivery codes assigned as the delivery progressed (**Figure 4**).

**Figure 4.**
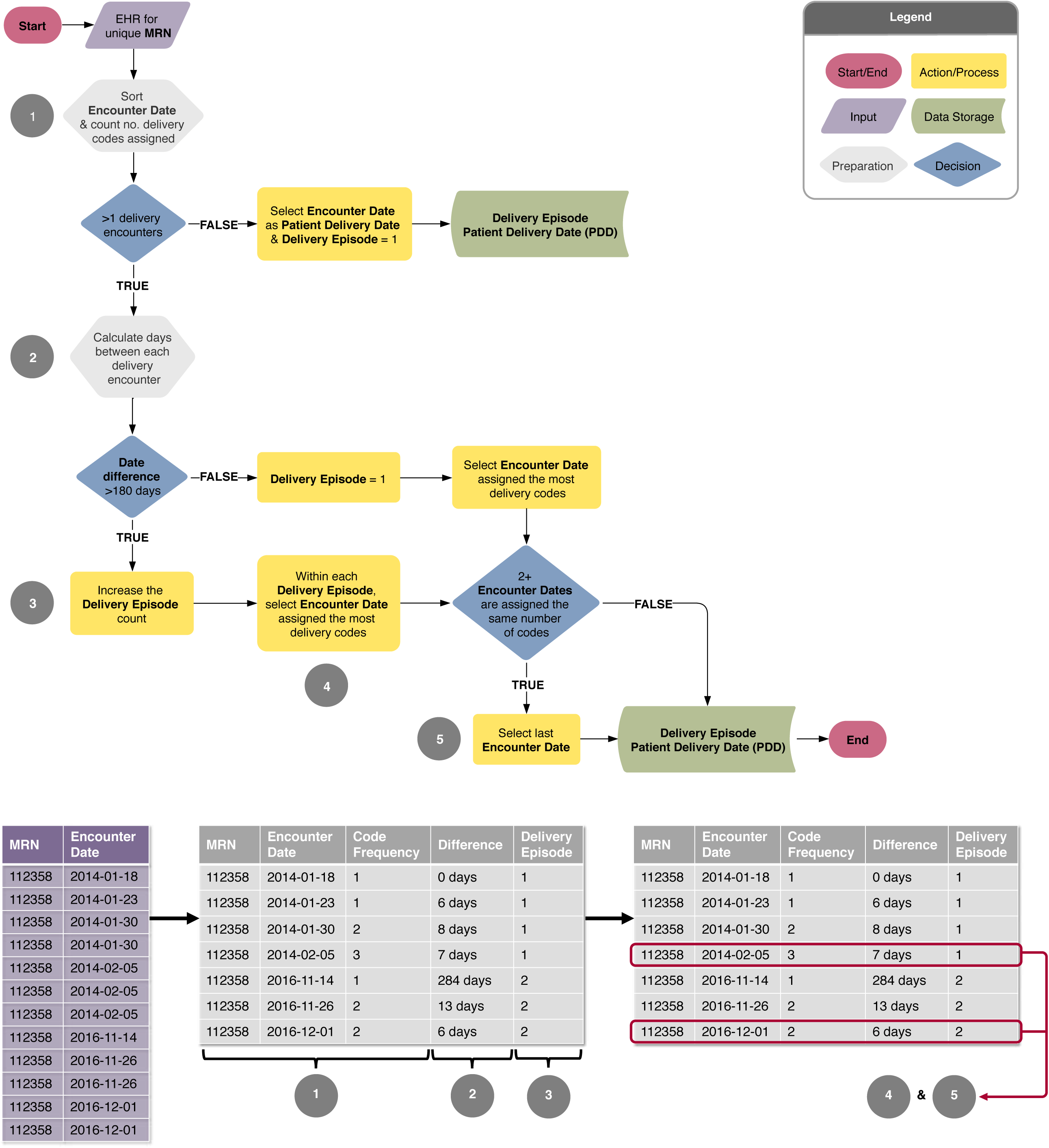
Overview and example of the MADDIE algorithm identifying separate delivery episodes using the delivery EHR of one fictitious patient. Abbreviations: *MRN*, Medical Record Number.

### 2.2 Algorithm Evaluation

The delivery episode algorithm was validated against an external record of births maintained by the Hospital of the University of Pennsylvania (HUP) within Penn Medicine, specifically by the Department of Obstetrics and Gynecology, to report vital statistics data to the state of Pennsylvania. For the purposes of validation, our dataset was limited to encounters at locations specifically labelled “HUP” or “Hospital of the University of Pennsylvania” to evaluate records relevant to the HUP obstetric provider locations. In addition, the birth log data were limited to the 2010-2017 time frame to match the time period of our dataset. The dataset validation subset contained 23,271 patients and 28,427 deliveries, and the birth log validation subset contained 26,260 patients and 31,646 birth records.

#### 2.2.1 Birth Log Gold Standard

The delivery date in the birth log was taken as the *true* delivery date and compared with the algorithm-selected PDD for validation. This birth log was maintained separately from the HER and was entered at the time of birth for all deliveries that occurred at HUP. Because it was independently maintained by the Department of Obstetrics and Gynecology, it represents an external source against which to validate our EHR algorithm. In addition to information on PDD, number of infants in womb (e.g., singleton, twins, triplets), the birth log contained additional delivery details such as infant outcome and birthweight. We used these additional details when manually reviewing any mismatches that occurred between MADDIE’s inferred PDD and the birth log’s true delivery date to ascertain any causes or systematic patterns for discrepancies when they occurred.

#### 2.2.2 Evaluating Patients with Single or Multiple Delivery Episodes

Patients with a one delivery were defined as having only one delivery episode within our dataset. Each patient ID and PDD were combined to create a unique delivery or pregnancy identification number, called the ‘pregnancy ID’. Patients with multiple deliveries were defined as having more than one delivery episode within our dataset. A pregnancy ID was created for each patient ID and PDD pair.

#### 2.2.3 Selecting Delivery Dates Within Episodes

In addition to evaluating MADDIE’s code frequency-based approach to selecting a delivery date, a random date-based approach was also evaluated, and the results of both compared. Instead of making assumptions about which delivery encounter would be the most likely PDD, the random date-based approach randomly selected an encounter date within each delivery episode.

#### 2.2.3.1 Code Frequency-Based Approach

For each patient, the time differences between the algorithm-selected PDD and all potential birth log delivery dates were calculated. PDDs with a difference of 30 days or less were considered ‘matches’ and evaluated using descriptive statistics. Those with a difference greater than 30 days were ‘mismatches’ and reviewed manually in the context of all the patient’s records to determine a reason for the mismatch (**Figure 5**).

**Figure 5.**
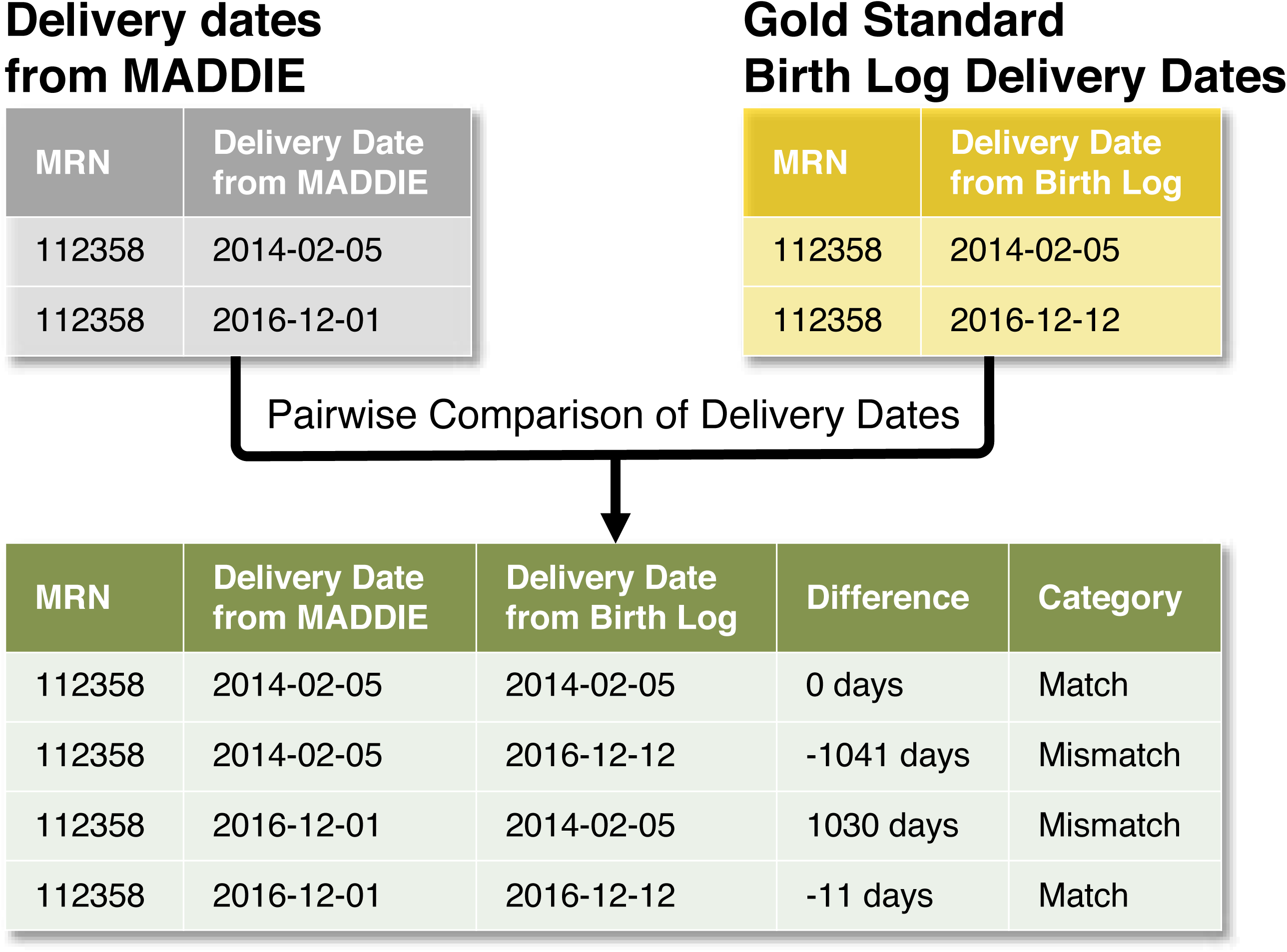
Example of validation of the MADDIE algorithm using the *code frequency-based approach* with the EHR and gold standard birth log delivery dates of one fictitious patient.

#### 2.2.3.2 Random Date-Based Approach

The random date-based approach randomly selected an encounter date within each delivery episode rather than make use of a heuristic (i.e. code frequency). The random date approach for selecting a PDD was repeated 100 times for each delivery episode, producing 100 random PDDs. The time difference between each of these 100 PDDs and all potential birth log delivery dates was calculated and relevant pairs were defined as those with the smallest time difference between them (i.e. one randomly selected PDD in 2011 should not be compared to a birth log delivery date in 2015 if a 2011 birth log delivery date is available). The average of these 100 relevant pairwise comparisons was then taken to produce one PDD and birth log delivery date time difference. The resulting time differences were then used to determine matches and mismatches as described for the code frequency-based approach (**Figure 6**).

**Figure 6.**
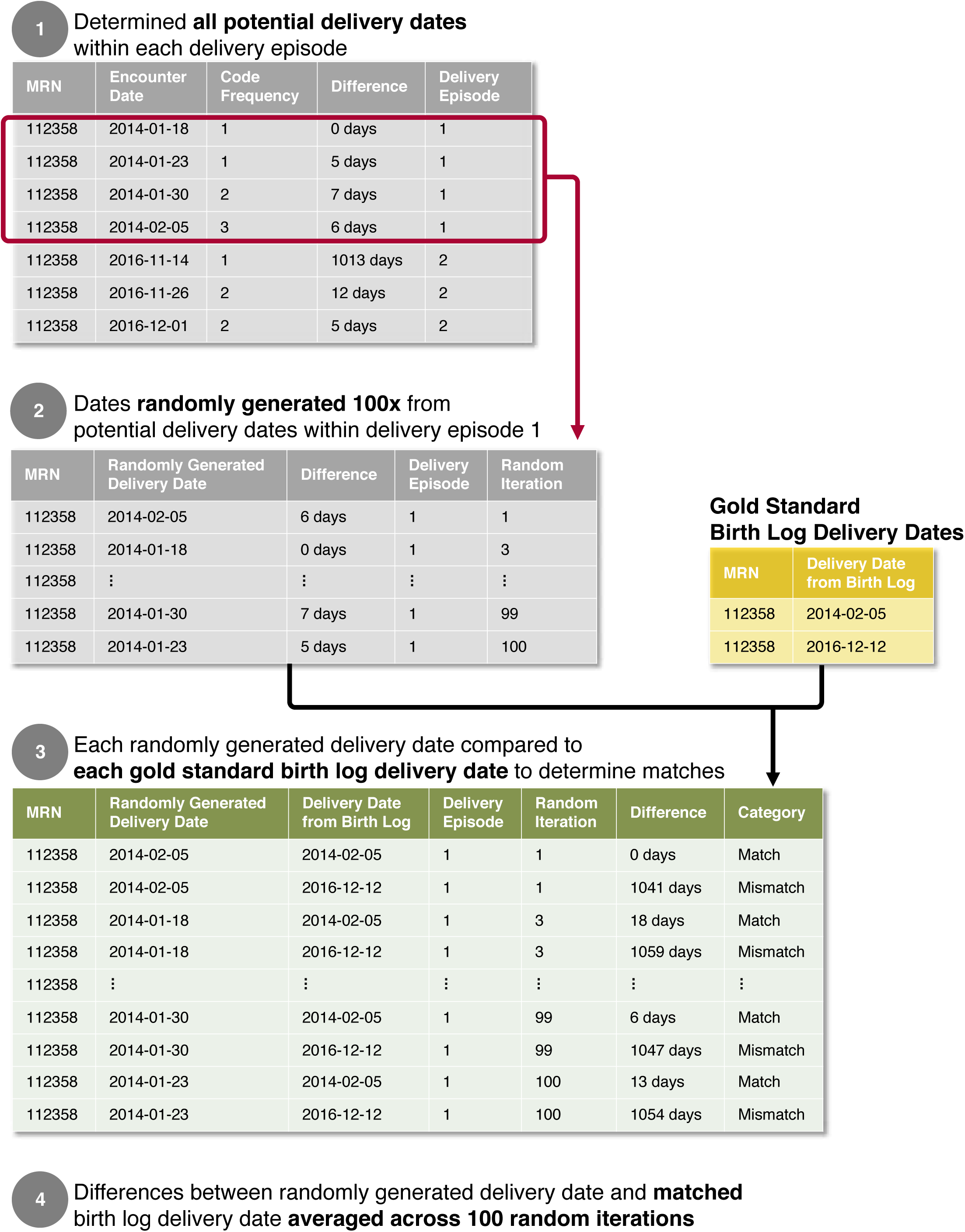
Example of validation of the MADDIE algorithm using the *random date-based approach* with the EHR and gold standard birth log delivery dates of one fictitious patient.

All code for MADDIE was implement in R (version 3.6.1) and EHR data was stored in a MySQL database. This study was approved by the Institutional Review Board of the University of Pennsylvania.

## 3. RESULTS

### 3.1 Dataset Characteristics

The algorithm identified 50,560 patients with delivery diagnoses or delivery procedures at HUP and a total of 63,334 pregnancies that resulted in delivery between 2010 and 2017. The average age at the time of delivery was 29.5 ± 6.1 years. The highest number of deliveries occurred in the 25-34 year age range making up 53.9% of deliveries. The most common race descriptions for patients included “black or African American” comprising 47.3% of deliveries, and “white” comprising 33.9% of deliveries (**Table 2**).

**Table 2.** Patient Characteristics of Patients that Delivered at Penn Medicine Identified by MADDIE.

| **Penn Medicine Population** | **MADDIE** |  |
| --- | --- | --- |
|  | **Patients (%)** | **Deliveries (%)** |
| Total population | 50560 (100) | 63334 (100) |
| **Age, years<sup>a</sup>** |  |  |
| < 18 | 1104 (2.2) | 1129 (1.8) |
| 18-24 | 11062 (21.9) | 12670 (20.0) |
| 25-34 | 28947 (57.25) | 34139 (53.9) |
| 35-44 | 13296 (26.3) | 15079 (23.8) |
| 45+ | 307 (0.6) | 317 (0.5) |
| Average: 29.5 ± 6.1 |  |  |
| **Race/Ethnicity<sup>b</sup>** |  |  |
| Black or African American | 23777 (47.1) | 29965 (47.3) |
| White | 17034 (33.7) | 21443 (33.9) |
| Hispanic | 4031 (8.0) | 4985 (7.9) |
| Asian | 3305 (6.5) | 4073 (6.4) |
| Other<sup>c</sup> | 2509 (5.0) | 2993 (4.7) |
| Native Hawaiian or other Pacific Islander | 75 (0.1) | 94 (0.1) |
| American Indian or Alaskan Native | 61 (0.1) | 81 (0.1) |

<sup>a</sup>Patient counts reflect the number of unique patients in each age bracket at the time of their delivery encounter. Patients with multiple deliveries are counted multiple times.
<sup>b</sup>Race/ethnicity descriptions are ‘non-Hispanic’ unless otherwise indicated.
<sup>c</sup>Other includes race description categories Other, Mixed, and Unknown.

The number of deliveries per patient at HUP ranged from one to seven with an average of 1.46 ± 0.67, and 62.2% of them being a first delivery within the dataset (**Table S1)**. The number of deliveries recorded was evenly distributed across all years except 2017 for which we had data for only part of the year. The average length of delivery episodes was 0.9 ± 10.5 days and the average time between delivery episodes for multiple deliveries was 2.6 ± 1.3 years.

### 3.2 Validation of the Algorithm Against the Gold Standard

In order to compare deliveries identified by MADDIE against those in the birth log, both patient populations were narrowed down to only those patients with encounter records specific to locations “HUP” or “Hospital of the University of Pennsylvania”. This resulted in validation populations of 23,271 unique patients with deliveries identified by MADDIE and 25,676 unique patients in the birth log. These validation patient populations were both found to have similar distributions of race/ethnicity with most patients identifying as “black or African American” in the MADDIE subset (66.0%) and the birth log subset (67.0%) (**Table 3**).

**Table 3.** Distribution of Race/Ethnicity Among Patients in Validation Sample for MADDIE and the Birth Log.

| Validation populations | MADDIE |  | Birth Log |
| --- | --- | --- | --- |
|  | Patients (%) | Deliveries (%) | Patients (%) |
| Total population | 23271 (100) | 28427 (100) | 25676 (100) |
| **Race/Ethnicity<sup>a</sup>** |  |  |  |
| Black or African American | 15363 (66.0) | 19004 (66.85) | 17193 (67) |
| White | 4383 (18.8) | 5301 (18.65) | 4595 (17.9) |
| Asian | 1539 (6.6) | 1812 (6.6) | 1614 (6.3) |
| Other/Not Available<sup>b</sup> | 1113 (4.8) | 1312 (4.6) | 1277 (4.9) |
| Hispanic | 904 (3.9) | 1044 (3.7) | 1034 (4.0) |
| Native Hawaiian or other Pacific Islander | 14 (0.1) | 17 (0.1) | 15 (0.1) |
| American Indian or Alaskan Native | 27 (0.1) | 34 (0.1) | 30 (0.1) |

<sup>a</sup>Race/ethnicity descriptions are ‘non-Hispanic’ unless otherwise indicated.
<sup>b</sup>Other includes race description categories Other, Mixed, and Unknown.
Not Available refers to refers to Birth Log records without a linked encounter record.

#### 3.2.1 Code Frequency-Based Approach Used in MADDIE vs. Random Date-Based Approach

When compared against the birth log, MADDIE was accurate within −0.68 ± 1.42 days for patients with one delivery and −0.52 ± 1.11 days (i.e. PDD is one half day earlier than the true delivery date) for those with two or more deliveries. Agreement between MADDIE and the birth log was 99.9% for cases with just one delivery and 98.5% for more than one delivery (i.e., more than one separate pregnancies).

The random-date based approach for inferring delivery dates was accurate within −0.71 ± 2.07 days for patients with one delivery and −0.57 ± 1.75 days for those with two or more deliveries.

Agreement between MADDIE and the birth log using this approach was 99.5% for one delivery and 98.0% for two or more delivery cases. The results obtained using MADDIE’s code-frequency-based approach were closer to the gold-standard birth log (−0.68 days) then the random date-based approach (−0.71 days) indicating that the code frequency-based approach is optimal. We used the code frequency-based approach in the MADDIE algorithm.

#### 3.2.2 MADDIE’s Overall Performance

To assess the performance of the MADDIE algorithm, we assessed its ability to accurately detect patients with deliveries occurring at HUP (the location with the birth log). MADDIE has a true positive rate, or **sensitivity**, of 89.6% and true negative rate, or **specificity**, of 99.8%. Its positive predictive value (PPV), or **precision**, was 98.8% and negative predictive value (NPV) was 98.5%. MADDIE was 98.5% **accurate** in identifying patients with deliveries at HUP and had an **F_1_ score** of 92.1% (**Table 4**).

**Table 4.**
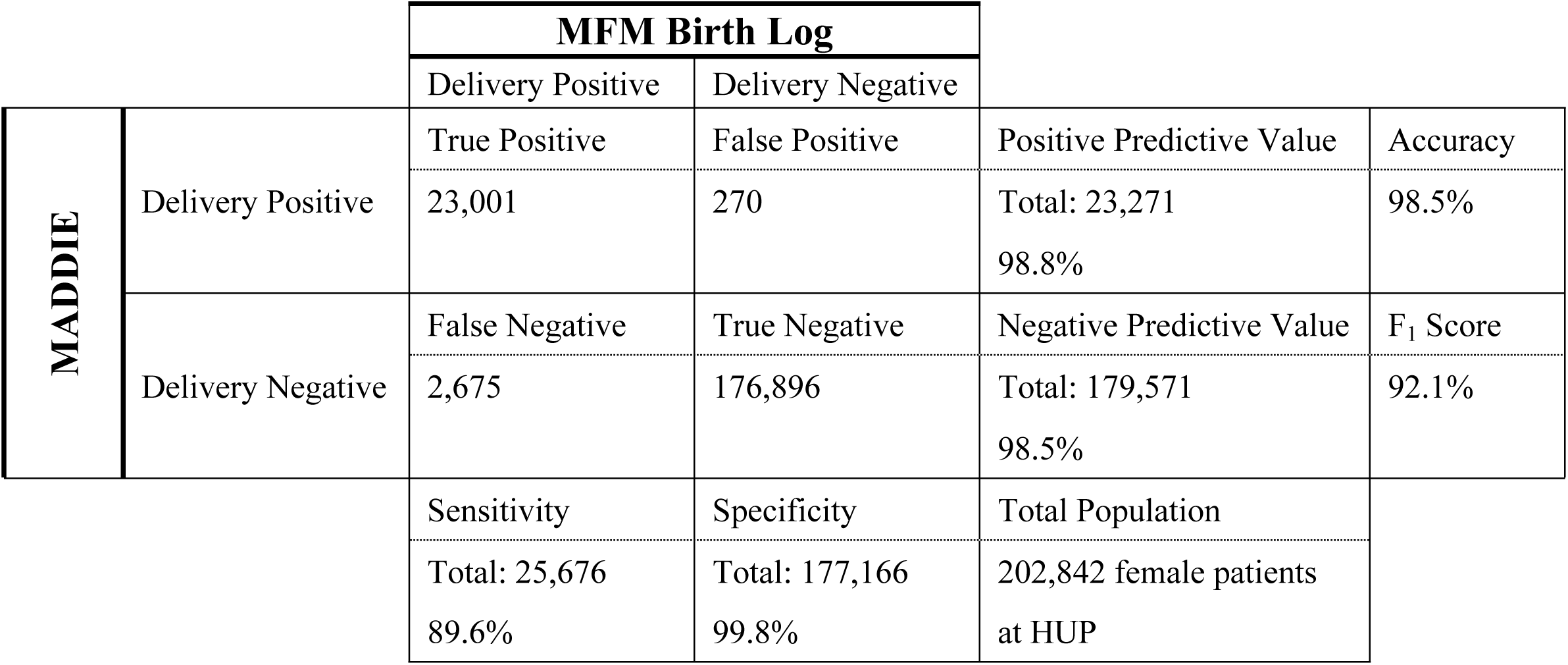
Performance Metrics for the Algorithm in Identifying Patients with Deliveries.

## 4. DISCUSSION

In this study, we developed the MADDIE algorithm to automatically infer delivery date information and delivery-specific details from medical records contained within the Penn Medicine EHRs. Overall MADDIE was 98.5% accurate with an F_1_ score of 92.1%. The difference between the MADDIE-determined delivery date based on the prediction algorithm and the independently maintained birth log at HUP (one of the hospitals within Penn Medicine) was on average half a day earlier. This is likely because the delivery encounter date is also the admission date, and patients sometimes delivery one to two days later.

### 4.1 MADDIE is Validated Against an Independently Maintained Birth Log and Not Clinical Code Review Used in Other Studies

Our study is the first study that develops an EHR algorithm that utilizes structured delivery codes alone and validates that delivery date algorithm using an independent birth log resource with known delivery dates as an external form of validation. This is a more robust validation approach relative to previously published methods that have mainly relied on manual review by trained abstractors of a random sample of the data,[18–20,27] or on comparing the number or rate of an identified delivery outcome (i.e. live birth, stillbirth) to official published national statistics data.[21,23] These prior manual reviews often involved clinician review of patient charts containing a variety of pregnancy-related codes, which is not as robust as using an independently maintained birth log for validation.

Furthermore, our gold standard contained specific birth details and was not dependent on a manual review that can be prone to biases. In addition, MADDIE does not discard valuable EHR data, such as repeat instances of delivery codes on any given encounter date, and rather utilizes this information to inform the selection process. Reasons for these repeat instances of delivery codes included delivery codes being assigned at different clinics, dates being assigned different delivery codes, or delivery codes being processed for billing and recorded in the patient’s EHR after the true delivery date. All of this information is valuable and helps to inform the selection of the accurate delivery date. The MADDIE algorithm was 98.5% accurate with an F_1_ score of 92.1% (**Table 4**). Therefore, MADDIE accurately detected patients with deliveries that were recorded in the birth log.

### 4.2 Utility of MADDIE to Advance Maternal Health Research in Large Clinical Datasets

MADDIE has the potential to facilitate investigation of prenatal exposures with pregnancy-related outcomes, particularly through the incorporation of additional features outside the scope of this study, such as medications. The major strength of MADDIE is its ability to distinguish delivery episodes from one another. Each patient’s timeline is broken into each distinct pregnancy that leads to a delivery. This allows researchers to study more than one delivery per patient and to adjust for patient-specific effects.

MADDIE is an improvement over other methods because it includes records from a large and racially/ethnically diverse patient population over multiple years from both inpatient and outpatient clinics. These characteristics allowed the dataset to capture less common clinical complications that provided an opportunity to address generalizability challenges when designing and refining the algorithm. In addition, MADDIE relies only on the structured elements from the EHR along with temporal information and the frequency of code usage to identify the patient delivery date, making it easily portable to other institutions and patient populations.

### 4.3 Limitations and Future Work

Delivery date discrepancies between MADDIE and the birth log were rare. In most cases these occurred between MADDIE and the gold standard because of early onset deliveries that required hospitalization for many days before the delivery occurred. This may be particularly relevant for centers such as HUP, as it is a tertiary care center where women with complicated pregnancies are admitted for observation and treatment prior to delivery. We speculate that in other, more, community-based centers, the algorithm may have even higher accuracy.

In some cases, a delivery was recorded in the birth log, but MADDIE did not detect it. This occurred because certain terminations and miscarriages were present in the birth log (our gold-standard), but these were not captured by MADDIE. The birth log contained records of deliveries that did not reach 20 weeks (i.e. reached 10 weeks gestation) and therefore may not have received a delivery code and these deliveries in the birth log also included some pregnancy terminations and miscarriages. We purposely did not include clinical codes for fetal loss, elective termination, induced abortion and other miscarriage codes in our MADDIE algorithm because we designed MADDIE to focus on deliveries, and we specifically used delivery codes typically assigned for deliveries occurring after 20 weeks of gestation. Therefore, while not capturing these earlier-stage pregnancy losses lowers MADDIE’s performance (because these early-stage pregnancies were listed in the birth log), we did not design MADDIE to uncover these early-stage pregnancy losses and therefore it did not uncover them. We see this as a strength of MADDIE as it found deliveries with at least 20 weeks gestation. Future work, involves developing a fetal loss and termination module as future work.

## 5. CONCLUSION

In this study, we developed the MADDIE algorithm to automatically infer delivery date information and delivery-specific details from medical records contained within the Penn Medicine EHRs. Overall MADDIE was 98.5% accurate with an F_1_ score of 92.1%. The MADDIE algorithm facilitates population-based studies of pregnant patients that are commonly underrepresented in clinical research. MADDIE allows for greater granularity in studying pregnancy episode-specific as well as delivery-specific associations using EHR data because it has the ability to distinguish patients having several distinct deliveries in the same EHR or clinical records system.

## CONTRIBUTORSHIP STATEMENT

Conceived Study Design: MRB

Developed methodology: SPC, MRB

Provided pertinent clinical advice to study problem: HHB, LDL

Wrote Paper: SPC, MRB

Reviewed, Edited, and Approved Final Manuscript: SPC, HHB, LDL, MRB

### Summary Points

- We designed an algorithm - MADDIE to extract patient delivery date information from EHR data
- MADDIE was 98.5% accurate with an F_1_ score of 92.1%
- We validated the algorithm against an independent gold-standard birth log

## Data Availability

All diagnosis and procedure code sets used in this analysis is included as a supplemental file with the publication of the paper. The patient-level data are not available for sharing due to privacy concerns, but relevant summary statistics are provided in the manuscript.

## CRediT authorship statement

**Mary Regina Boland:** Conceptualization, Methodology, Software, Validation, Formal analysis, Investigation, Resources, Data Curation, Writing – Review & Editing, Supervision, Project administration, Funding acquisition. **Silvia Canelón:** Methodology, Software, Validation, Formal analysis, Investigation, Data Curation, Writing – Original draft preparation, Writing – Reviewing & Editing, Visualization. **Heather Burris:** Writing – Review & Editing. **Lisa Levine:** Writing – Review & Editing.

## Supplemental Tables for

### Table of Contents

**Table S1.**
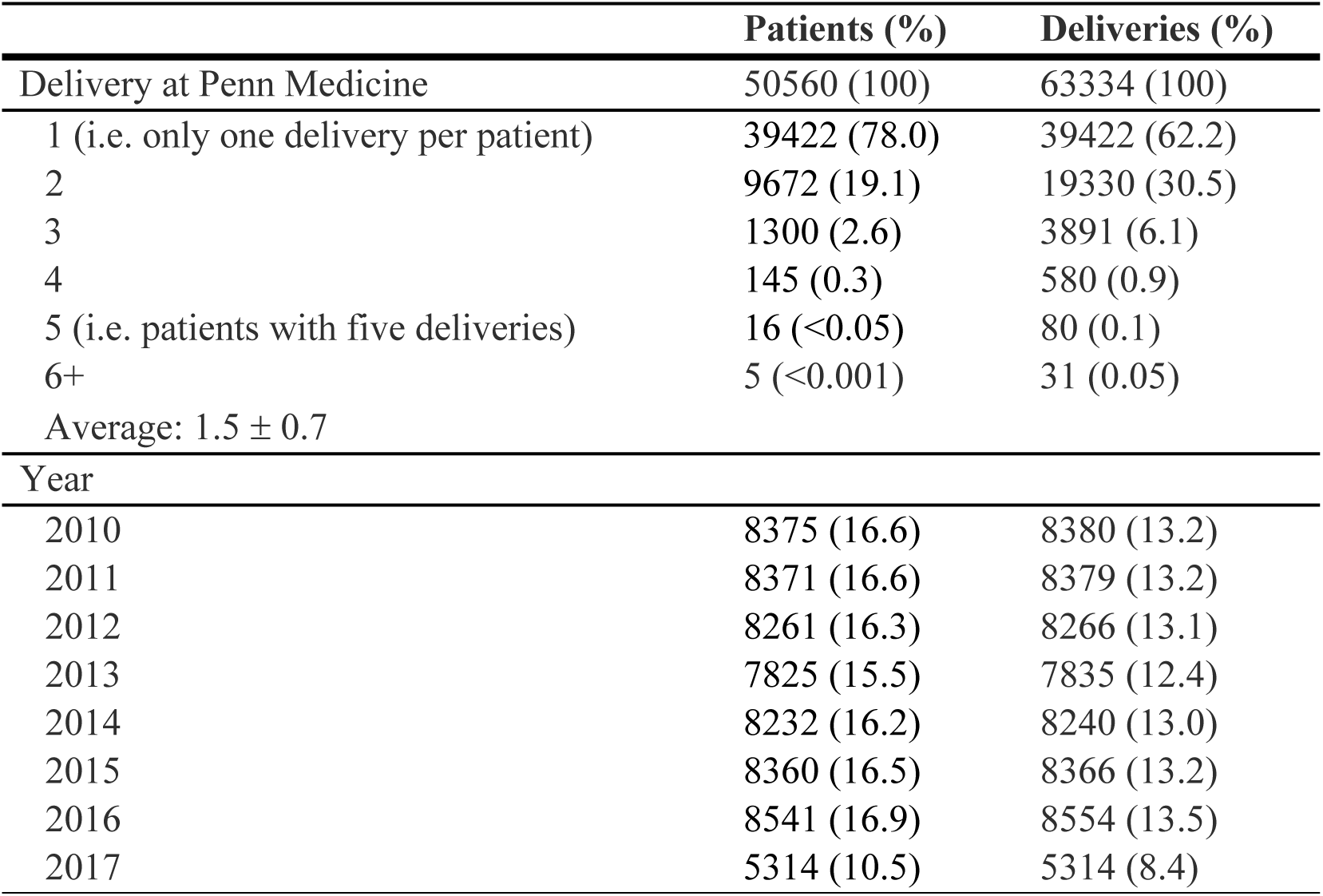
The Number of Deliveries per Year and per Patient Identified by MADDIE at Penn Medicine.

|  | **Patients (%)** | **Deliveries (%)** |
| --- | --- | --- |
| Delivery at Penn Medicine | 50560 (100) | 63334 (100) |
| 1 (i.e. only one delivery per patient) | 39422 (78.0) | 39422 (62.2) |
| 2 | 9672 (19.1) | 19330 (30.5) |
| 3 | 1300 (2.6) | 3891 (6.1) |
| 4 | 145 (0.3) | 580 (0.9) |
| 5 (i.e. patients with five deliveries) | 16 (<0.05) | 80 (0.1) |
| 6+ | 5 (<0.001) | 31 (0.05) |
| Average: $1.5 \pm 0.7$ | | |
| Year |  |  |
| 2010 | 8375 (16.6) | 8380 (13.2) |
| 2011 | 8371 (16.6) | 8379 (13.2) |
| 2012 | 8261 (16.3) | 8266 (13.1) |
| 2013 | 7825 (15.5) | 7835 (12.4) |
| 2014 | 8232 (16.2) | 8240 (13.0) |
| 2015 | 8360 (16.5) | 8366 (13.2) |
| 2016 | 8541 (16.9) | 8554 (13.5) |
| 2017 | 5314 (10.5) | 5314 (8.4) |

